# Data use practices and challenges for maternal and child health decision-making in tribal primary health centres in Andhra Pradesh, India

**DOI:** 10.64898/2026.03.29.26349634

**Authors:** Arun Mitra, Gurukartick Jayaraman, Bhavana Ondopu, Santosh Kumar Malisetty, Raghu Niranjan, Shaheen Shaik, Biju Soman, Rakhal Gaitonde, Tarun Bhatnagar, Engelbert Niehaus, Sajinkumar K.S, Adrija Roy

## Abstract

**Background:** Primary health centres in tribal areas of India collect large volumes of maternal and child health (MCH) data through routine health information systems, yet this data rarely informs local clinical or programmatic decision-making. The gap between data collection and data use in tribal settings, where health disparities are most acute, remains poorly documented from the perspective of frontline decision-makers.

**Methods:** We conducted a qualitative study embedded in the diagnostic phase of an Action Research project in three tribal primary health centres under the Integrated Tribal Development Agency (ITDA), Rampachodavaram, Alluri Sitharama Raju District, Andhra Pradesh. Eight key informant interviews were conducted with medical officers (n=5), a district programme officer (n=1), and data entry operators (n=2). Participant observation at weekly convergence meetings and document review of registers and reports supplemented interview data. Transcripts were independently coded by two analysts using Braun and Clarke’s reflexive thematic analysis.

**Findings:** Three interconnected domains emerged. First, local MCH decision-makers needed individual-level, geographically disaggregated, prospective information to plan outreach and follow-up, but formal systems provided only retrospective aggregate statistics. Second, three structural constraints prevented formal systems from meeting these needs: digital infrastructure designed for connected settings, upward data flows with no local feedback, and a single-point-of-access governance vulnerability where one data entry operator’s mobile phone controlled portal authentication for all facilities in the jurisdiction. Third, decision-makers constructed four complementary information practices (WhatsApp networks, self-built tracking tools, cross-sectoral convergence meetings, and reliance on intermediary-consolidated reports) to bridge the gap.

**Interpretation:** Complementary information practices are expressions of local ingenuity under structural constraint, not system failures. MCH digital health reform should map and strengthen these practices rather than bypass them. Authentication governance in low-connectivity tribal settings requires urgent policy attention.

**Funding:** DST-SEED Fellowship.

**Research in Context:** *Evidence before this study.:* Routine health information system (RHIS) literature has extensively documented the gap between data collection and data use in low- and middle-income countries. Systematic reviews identify organisational, technical, and behavioural determinants of data use at the facility level. However, qualitative evidence specifically addressing MCH decision-making in tribal or indigenous health settings in India is limited. Informal information practices that local decision-makers construct to compensate for formal system failures, including WhatsApp-based clinical communication, self-built digital tracking tools, and human data intermediaries, are largely undocumented in the peer-reviewed literature. The governance consequences of centralised OTP-based authentication credential registration in low-connectivity settings have not previously been described.

*Added value of this study.:* This study provides the first systematic qualitative account of how local MCH decision-makers navigate between formal health information systems and self-constructed information practices in a tribal district of Andhra Pradesh. It reveals a structural single-point-of-access vulnerability at the sub-district level, where a single data entry operator’s registered mobile number controls portal authentication for all primary health centres in the jurisdiction. It applies the Open Data Institute’s data ecosystem framework to surface complementary information practices as soft value exchanges alongside formal data flows.

*Implications of all the available evidence.:* MCH digital health interventions should conduct participatory diagnostics of local information ecosystems before tool deployment. Data access governance at the sub-district level, including authentication credential registration policy, requires reform to prevent single points of failure. Complementary information practices are latent MCH infrastructure that should be formalised and strengthened, not replaced by new systems that ignore existing local capacity.

## 1 Introduction

Primary health centres in tribal India occupy a paradoxical position in the health information landscape. They are sites of intensive data collection, with frontline health workers recording maternal and child health service events across multiple platforms, registers, and reporting formats. Yet the same facilities lack access to their own data in forms that support local clinical or programmatic decision-making. Data flows upward through administrative hierarchies to district and state levels; it rarely returns in forms that enable local action.

This is not a new observation. The gap between data availability and data use has been documented across low- and middle-income country settings, with systematic reviews identifying organisational, technical, and behavioural determinants (1–3). What remains less well understood is how this gap operates in specific institutional contexts, particularly in tribal areas where geographic isolation, governance complexity, and population characteristics create distinctive challenges for health information systems.

India’s tribal populations, constituting 8.6% of the national population, experience persistent health disparities including higher maternal and infant mortality, lower immunisation coverage, and reduced access to quality care (4). In Andhra Pradesh, the Integrated Tribal Development Agency (ITDA) model introduces an additional governance layer between the district health administration and primary health centres, creating distinctive data flows and accountability structures. The recent formation of Alluri Sitharama Raju (ASR) District in April 2022, carved from the tribal portions of East Godavari and Visakhapatnam, further reconfigured administrative arrangements during the study period.

The formal health information systems serving these areas were not designed with tribal primary health centres as the primary user (5,6). The Reproductive and Child Health (RCH) Portal, the Health Management Information System (HMIS), and associated digital platforms were built for upward reporting: aggregating facility-level data for district, state, and national monitoring (7). Their interfaces prioritise data entry over data retrieval. Their outputs serve supervisory review, not frontline decision support. As one medical officer in the study described it:

> “Whatever the apps were there, they were just to enter the field data of what they have done. Most of it was like this only and not to be reviewed by the medical officer to use for the field level. It was like only one way.” (MO, Boduluru)

This upward accountability orientation is a structural design feature, not an individual capacity problem. Understanding how it operates at the facility level, what information needs remain unmet, and what practices local decision-makers construct in response, is essential for designing MCH interventions that are contextually appropriate rather than generically imposed.

This study aimed to understand what information local MCH decision-makers need, what they currently do with available data, and what structural conditions prevent effective data use for local MCH action. The findings were generated during the diagnostic phase of a participatory data science project, preceding any intervention design.

## 2 Methods

### 2.1 Study design

This qualitative study was embedded in the diagnostic phase of an Action Research project following the cyclic framework of Susman and Evered (8). The diagnostic phase sought to surface contextual realities of MCH data use before any intervention was designed. The study adopted an interpretivist orientation, recognising that data practices are shaped by institutional structures, local meanings, and relational dynamics that quantitative approaches alone cannot capture.

### 2.2 Setting and participants

The study was conducted in three primary health centres under ITDA Rampachodavaram, Alluri Sitharama Raju District, Andhra Pradesh (Figure 1). PHC Boduluru (Maredumilli mandal), PHC Gangavaram (Gangavaram mandal), and PHC Vadapalli (Rampachodavaram mandal) were purposively selected to represent variation in geographic accessibility, connectivity infrastructure, and tribal population composition (Table 1).

**Figure 1:**
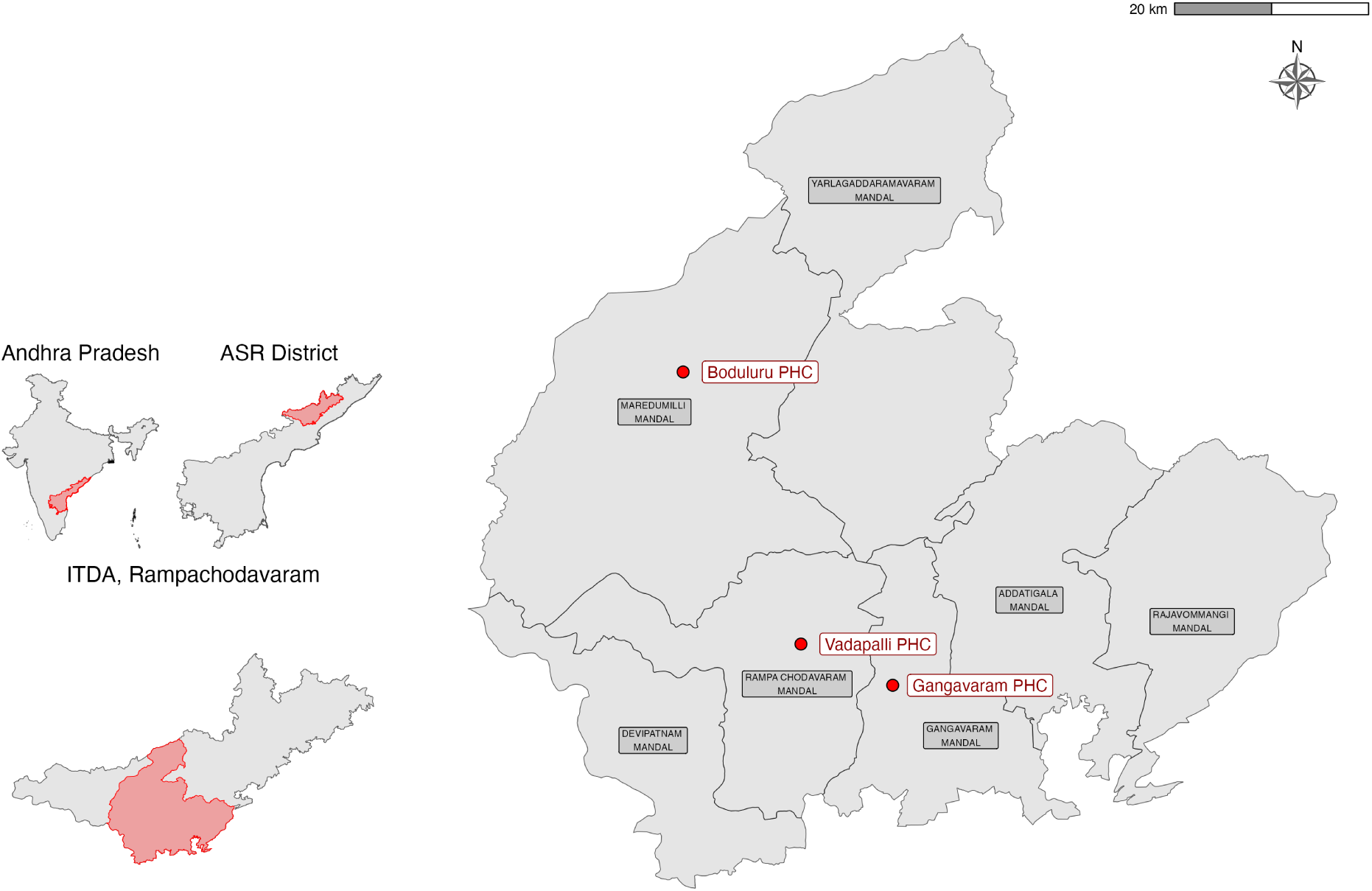
Geographical location of the three study PHCs under ITDA Rampachodavaram, Alluri Sitharama Raju District, Andhra Pradesh, India. Inset shows the position of ASR District within Andhra Pradesh.

**Table 1:** Study site characteristics. Mandal-level data from ITDA Rampachodavaram administrative records. PVTG = Particularly Vulnerable Tribal Group.

| Characteristic | Boduluru | Gangavaram | Vadapalli |
| --- | --- | --- | --- |
| Mandal | Maredumilli | Gangavaram | Rampachodavaram |
| Mandal area (km<sup>2</sup>) | 913.96 | 612.71 | 610.30 |
| Forest cover (%) | 81.6 | 69.4 | 65.1 |
| Terrain | Hilly, forested | Moderately hilly | Hilly, remote |
| Road connectivity | Limited | Moderate | Limited |
| Mobile network | Inconsistent | Moderate | Inconsistent |
| Village Secretariats (n) | 8 | 10 | 16 |
| Panchayats (n) | 12 | 17 | 19 |
| Villages (n) | 68 | 55 | 77 |
| Habitations (n) | 113 | 79 | 102 |
| Households (n) | 4,801 | 7,550 | 10,554 |
| Scheduled Tribe population (%) | 93.3 | 67.2 | 79.3 |
| PVTG population (% of ST) | 39.8 | 1.3 | 2.4 |
| MO community status | Non-local | Community member | Non-local |

Participants included medical officers (MOs), who serve as the primary local MCH decision-makers at the PHC level; auxiliary nurse midwives (ANMs), who generate field-level data and deliver sub-centre services; data entry operators (DEOs), who bridge paper records and digital systems; and ITDA programme staff including the Multipurpose Health Data Entry Operator (MPHDEO) at the sub-district level.

Eight key informant interviews were conducted. The study population was bounded by institutional structure (three PHCs under one ITDA), and all medical officers within the jurisdiction were included. Sampling aimed for comprehensive coverage of the decision-making chain rather than statistical saturation; the three data collection methods (interviews, observation, document review) provided triangulation across sources (Table 2).

**Table 2:**
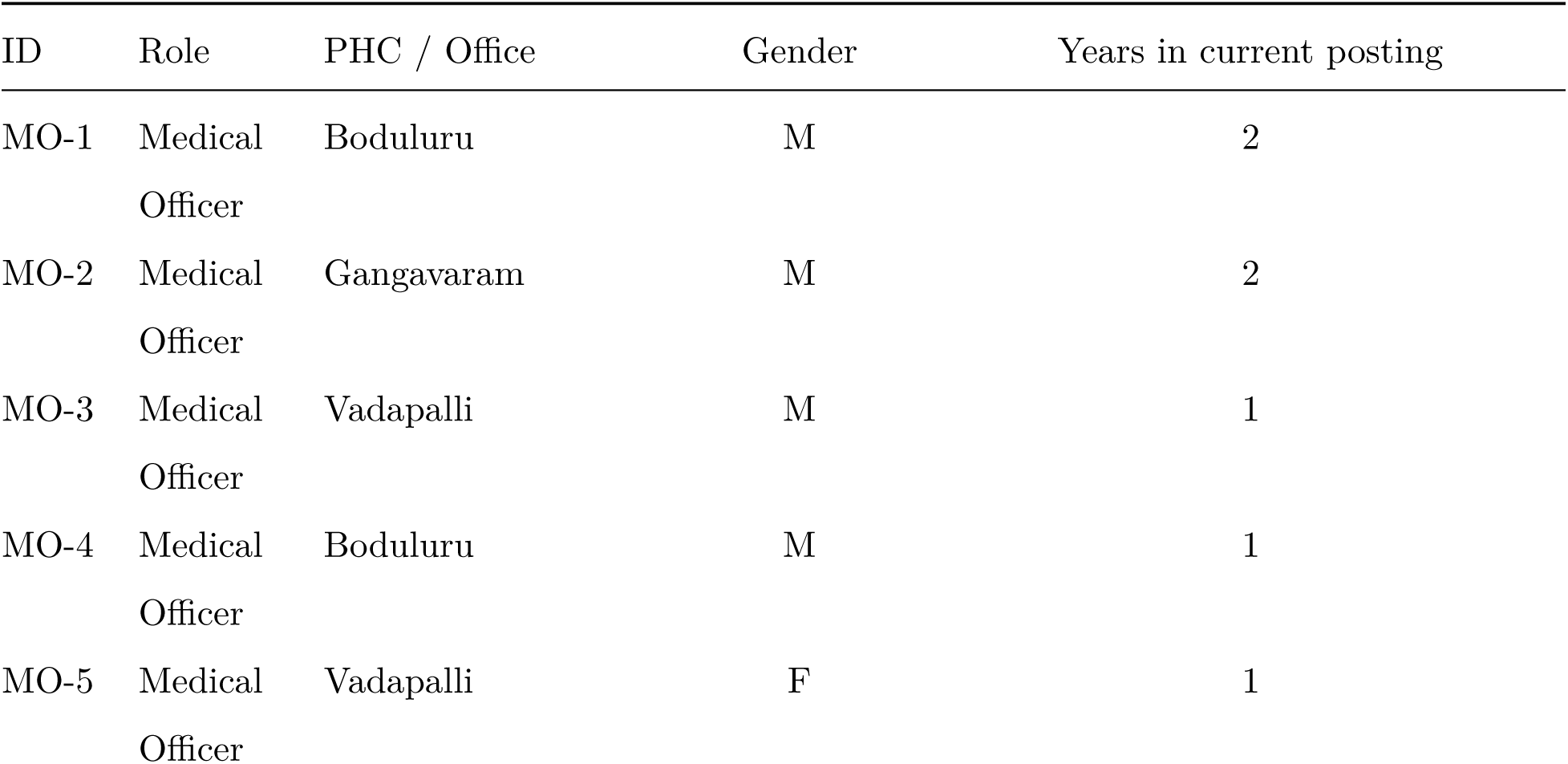
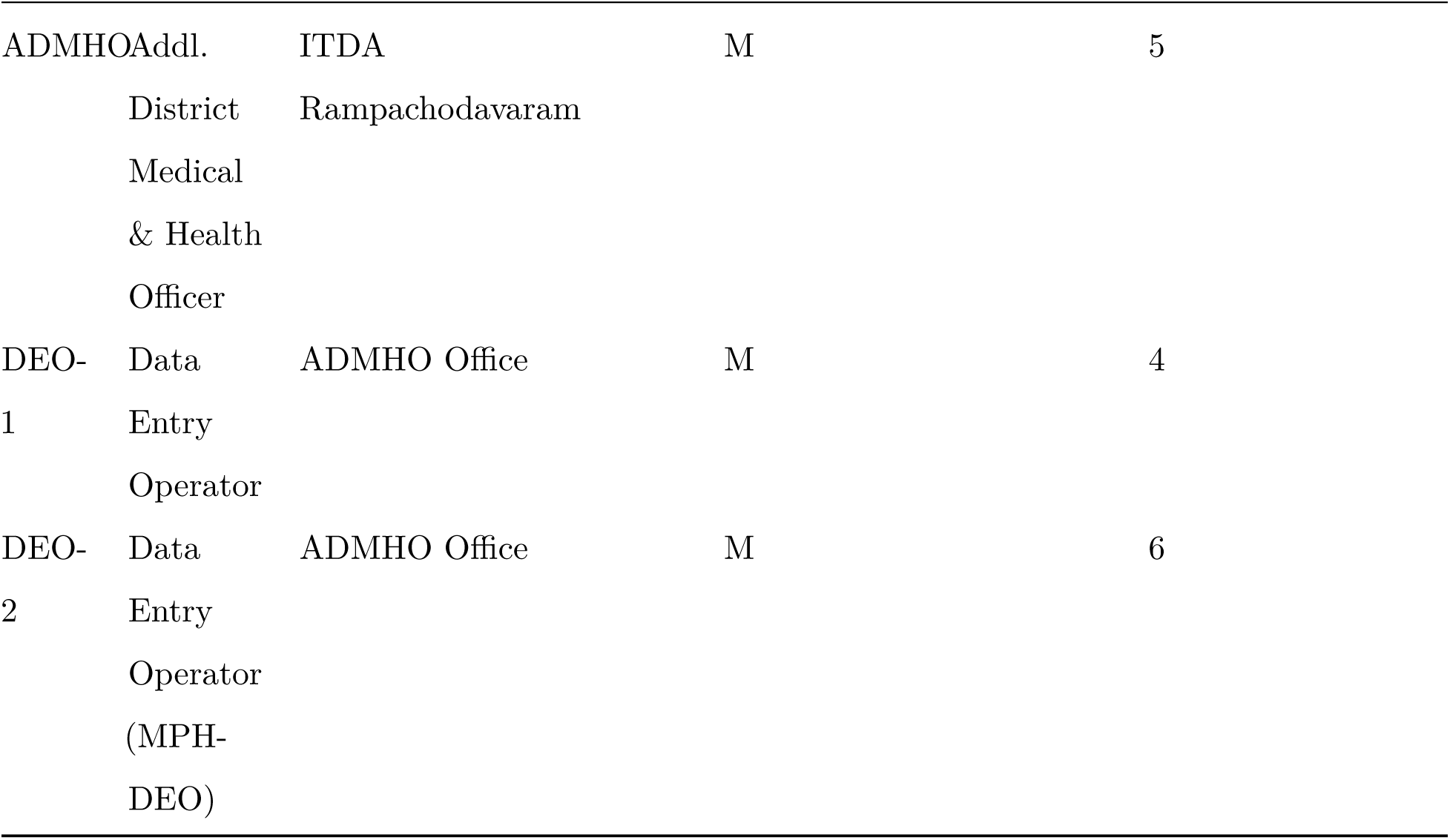
Participant characteristics.

### 2.3 Data collection

Data were collected between August 2023 and January 2024 through three methods. Eight key informant interviews (KIIs) were conducted: five with medical officers across the three PHCs, one with the Additional District Medical and Health Officer (ADMHO) at ITDA Rampachodavaram, and two with data entry operators at the ADMHO office. Interviews followed a semi-structured guide covering MCH data use practices, decision-making routines, system experiences, and information needs. Interviews were conducted in English and Telugu, audio-recorded with consent, and transcribed verbatim.

Participant observation was conducted at weekly cross-sectoral convergence meetings (Thursdays at all three PHCs), where medical officers, ANMs, ASHAs, and Anganwadi workers convene to discuss service delivery and data issues. Observation notes documented data-related discussions, system access practices, and informal information exchanges.

Document review encompassed paper registers (ANC, immunisation, delivery), Mother and Child Protection (MCP) cards, monthly HMIS reports, informal consolidated reports circulated via WhatsApp, and personal tracking tools maintained by individual medical officers.

### 2.4 Analysis

Transcripts were analysed using Braun and Clarke’s six-phase reflexive thematic analysis (9). Two analysts (AM and GJ) independently coded the transcripts using both inductive and deductive approaches. Deductive codes were drawn from existing RHIS and data utilisation literature; inductive codes allowed themes to emerge from the data beyond existing frameworks. Coding discrepancies were resolved through deliberation and reference to original transcripts. Emergent themes were organised into three analytical domains: information needs, structural constraints on formal data systems, and complementary information practices. Member-checking with medical officers was conducted to validate interpretive accuracy. The Open Data Institute’s data ecosystem framework was applied to distinguish formal data flows (value exchanges through official channels) from soft value exchanges (flows of insights, knowledge, and feedback operating outside formal infrastructure) (10).

### 2.5 Positionality

The lead researcher (AM) is a member of the tribal community in the ITDA Rampachodavaram area. This insider status facilitated research access, trust-building, and contextual interpretation, while also requiring reflexive attention to assumptions derived from personal experience. Strategies to manage insider effects included regular supervisor debriefs, systematic attention to disconfirming evidence, and the participatory orientation of the research itself, in which stakeholders were partners in sense-making rather than merely data sources.

### 2.6 Ethics

The study received ethical approval from the Institutional Ethics Committee, Sree Chitra Tirunal Institute for Medical Sciences and Technology (Protocol No. SCT/IEC/2047/MAY/2023). ITDA Rampachodavaram community research protocols were followed. Written informed consent was obtained from all participants. The CONSIDER statement for research involving Indigenous peoples informed the ethical framework (11). Participant names are anonymised throughout; the MPHDEO is referenced by institutional function, not by name.

## 3 Findings

Three interconnected domains emerged from the analysis: what local MCH decision-makers need but cannot access from formal systems; the structural constraints that explain why; and the complementary information practices they construct in response.

### 3.1 What local MCH decision-makers need: actionable individual intelligence

Across all three PHCs, a consistent set of unmet information needs emerged. Notably, no medical officer expressed a need for more data. Frustration was uniformly about access, timeliness, and granularity of data that already existed within the system but was inaccessible at the point of decision-making.

The most consistently expressed need was for individual-level line lists tied to geography. Medical officers needed to know which specific beneficiary, in which village or Sachivalayam (village secretariat), was overdue for which MCH service:

> “If they say 5 pending, there is no use of someone telling me you have 5 pending. They have to tell me that you have 5 pending in these and these Sachivalayams. So these and these are to be completed.” (MO, Vadapalli)

This “names over numbers” imperative reflected the operational reality of PHC-level decision-making. Aggregate coverage percentages are meaningful for district-level monitoring but offer no guidance for a medical officer deciding which villages to prioritise for outreach or which ANMs require supervision. ANMs expressed the same need from the field perspective:

> “They showed the percentage. Those who showed the daily, they showed the percentage. But I don’t know who it is.” (ANM, Gangavaram)

Decision-makers also wanted prospective rather than retrospective information. Existing systems reported what had been missed; medical officers wanted systems that flagged what needed to be done:

> “Instead of saying ‘you didn’t do this’, it would be better to say ‘this should be done’. It would be nice to say ‘here it is’ before saying ‘here it is, you missed it’.” (MO, Boduluru)

These information needs were grounded in concrete clinical workflows: planning immunisation schedules, tracking ANC attendance, identifying postnatal follow-up, and monitoring high-risk pregnancies. Medical officers articulated a clear desire for data that could be acted upon immediately:

> “If I have that data in my hand ready now whenever I want it… I could keep that information ready and pass that information not immediately but much faster.” (MO, Boduluru)

The information gap was therefore not about data availability. The RCH Portal contained individual-level records for every registered beneficiary. The gap was between what the system stored and what it made accessible to the people responsible for acting on it.

### 3.2 Structural constraints on formal MCH data use

Three categories of structural constraint prevented formal systems from meeting the information needs described above.

#### 3.2.1 Digital infrastructure designed for connected settings

All three PHCs experienced persistent connectivity failures that rendered portal-based systems unreliable for routine data access. The 2024 introduction of OTP-based authentication, designed to improve data security, compounded access problems in low-connectivity tribal settings. Authentication required receiving a one-time password via SMS on a registered mobile number, a process that assumed reliable mobile network coverage:

> “If everyone wants to bring everyone to the signal area, everyone… by the time you click the OTP, it will be completely passed. It takes about 20 minutes to complete it in one go.” (ANM, Gangavaram)

> “We don’t have signal here. If we have to attend [meetings], we will have to go all the way till Ramachoudavaram or at least Bandupalli.” (MO, Vadapalli)

The proliferation of digital tools further compounded the burden (12,13). Staff reported navigating dozens of platforms with distinct login credentials:

> “36 apps. Single app, different IDs and passwords.” (Staff, Gangavaram)

Earlier system configurations had permitted broader access at lower administrative cost. The shift to per-PHC logins with individual OTP requirements represented a security improvement that, in this context, functioned as an access barrier:

> “Earlier it was a sub-district level, the entire division’s line listing we could get through a single ID and password. But now it is each PHC, a separate login ID, a separate password, as well as a separate person’s registered mobile number for the OTP.” (MO, Vadapalli)

#### 3.2.2 Upward data flows with no local return

Formal systems were structured as upward reporting pipelines. Data entered at the facility level flowed to district and state levels for monitoring and programme management. No mechanism returned actionable information to the facilities that generated it:

> “At the end of the day, they only see the results. They don’t know how we are doing.” (MO, Vadapalli)

Targets set at the state level compounded the structural position of local decision-makers. Urban benchmarks were applied to facilities operating under fundamentally different resource constraints:

> “At the state level, they are giving targets to achieve by comparing these tribal PHCs with urban PHCs… But resources are less compared to the urban PHCs, and the targets are the same.” (MO, Boduluru)

The absence of a feedback loop meant that data submission functioned as an end in itself. Medical officers were accountable for entering data but received no analytical return on the time invested. Data served surveillance and accountability, not local learning.

#### 3.2.3 The MPHDEO as single point of data access

A critical structural finding emerged across all three PHCs. The ITDA Rampachodavaram’s institutional login credentials for the RCH Portal were linked to the mobile phone of a single Multipurpose Health Data Entry Operator (MPHDEO) at the sub-district level. Because OTP-based authentication required physical possession of this registered mobile device, the MPHDEO became the sole custodian of portal access across all PHCs in the jurisdiction.

Local decision-makers at Boduluru, Gangavaram, and Vadapalli were structurally dependent on this single individual to retrieve any data from the formal system:

> “That report is helpful… It will take a lot of time [without it]. We don’t have access to everything that [the MPHDEO] keeps.” (MO, Vadapalli)

This arrangement was not the result of deliberate policy design but an unintended consequence of how authentication credentials were registered at system rollout. It represents a structural vulnerability in MCH data governance: concentrating access rights in a single non-clinical actor at the sub-district level, effectively creating an information bottleneck between the formal data system and the local decision-makers it was designed to serve. When this individual is transferred or unavailable, institutional data access is severed entirely.

### 3.3 Complementary information practices: local decision-makers filling the gap

Faced with formal systems that could not deliver the information they needed, local MCH decision-makers constructed their own practices. Applying the Open Data Institute’s data ecosystem framework, these constitute soft value exchanges: flows of insights, knowledge, and feedback operating outside formal data infrastructure (10). Four complementary information practices were identified across the three PHCs (Table 3).

**Table 3:** Complementary information practices identified across study PHCs.

| Practice | Mechanism | MCH function | Spectrum position |
| --- | --- | --- | --- |
| WhatsApp-based case communication | Case photos, beneficiary updates, alerts shared in MO-ANM-ASHA groups | Real-time individual-level MCH visibility | Complementary |
| Self-built tracking tools | Google Sheets immunisation tracker populated by field staff | Due-list functionality replacing portal | Substitutive |
| Convergence meetings | Weekly in-person cross-sectoral meetings resolving data discrepancies | Data triangulation across registers | Complementary |
| MPHDEO consolidated reports | Sub-district DEO synthesises data across facilities, distributes via WhatsApp | Aggregate MCH intelligence for PHC planning | Compensatory |

#### 3.3.1 WhatsApp as real-time case communication

ANMs and ASHAs shared case photographs, beneficiary updates, and urgent alerts through WhatsApp groups that included medical officers (14). This informal channel delivered real-time individual-level MCH visibility that the RCH Portal could not:

> “In WhatsApp and WhatsApp. That’s it. So because they share it by WhatsApp.” (MO, Boduluru)

Medical officers envisioned formalising this channel rather than replacing it:

> “WhatsApp is there, that’s easy. If there is a dashboard that can give you the alert also through WhatsApp, it would be like a superpower in my hands.” (MO, Boduluru)

#### 3.3.2 Self-built tracking tools for MCH monitoring

One medical officer independently constructed a Google Sheets immunisation tracker, assigning field staff to populate it and reviewing it by date (15). This replicated the due-list functionality that formal portals were designed but failed to provide:

> “I Google Sheet, I almost all of it. I will simply do it and form it. I will put it on Google Sheets. They will fill it in there. I will check date-wise.” (MO, Gangavaram)

#### 3.3.3 Thursday cross-sectoral convergence meetings

One PHC institutionalised a weekly in-person meeting bringing together ANMs, ASHAs, and Anganwadi workers to resolve data discrepancies, address system access problems, and triangulate MCH information across registers. The meeting was explicitly designed around the connectivity constraint:

> “Every Thursday, since some people don’t have signal areas, come here every Thursday. If you have any doubts, we can sort them out here.” (MO, Gangavaram)

#### 3.3.4 Reliance on the MPHDEO’s consolidated reports

Given the structural access barrier described above, all three PHCs depended on consolidated reports prepared by the MPHDEO, who synthesised data across facilities and distributed summaries via WhatsApp as and when needed or requested by PHC staff. As noted above, one medical officer described these reports as the most valued information product in the local ecosystem. What began as a structural constraint had been partially converted into a complementary practice, with the MPHDEO functioning as a human data intermediary.

These four practices range from complementary (convergence meetings, which sit alongside formal reporting) to substitutive (Google Sheets, which replaces formal portal functionality entirely). They are not workarounds in the pejorative sense. They are evidence of deliberate local problem-solving under structural constraint, constituting an informal MCH information ecosystem that is parallel to, and frequently more responsive than, the formal HIS.

Figure 2 summarises the relationships between the three analytical domains: unmet information needs drive the emergence of complementary practices, mediated by the structural constraints that prevent formal systems from meeting those needs.

**Figure 2:**
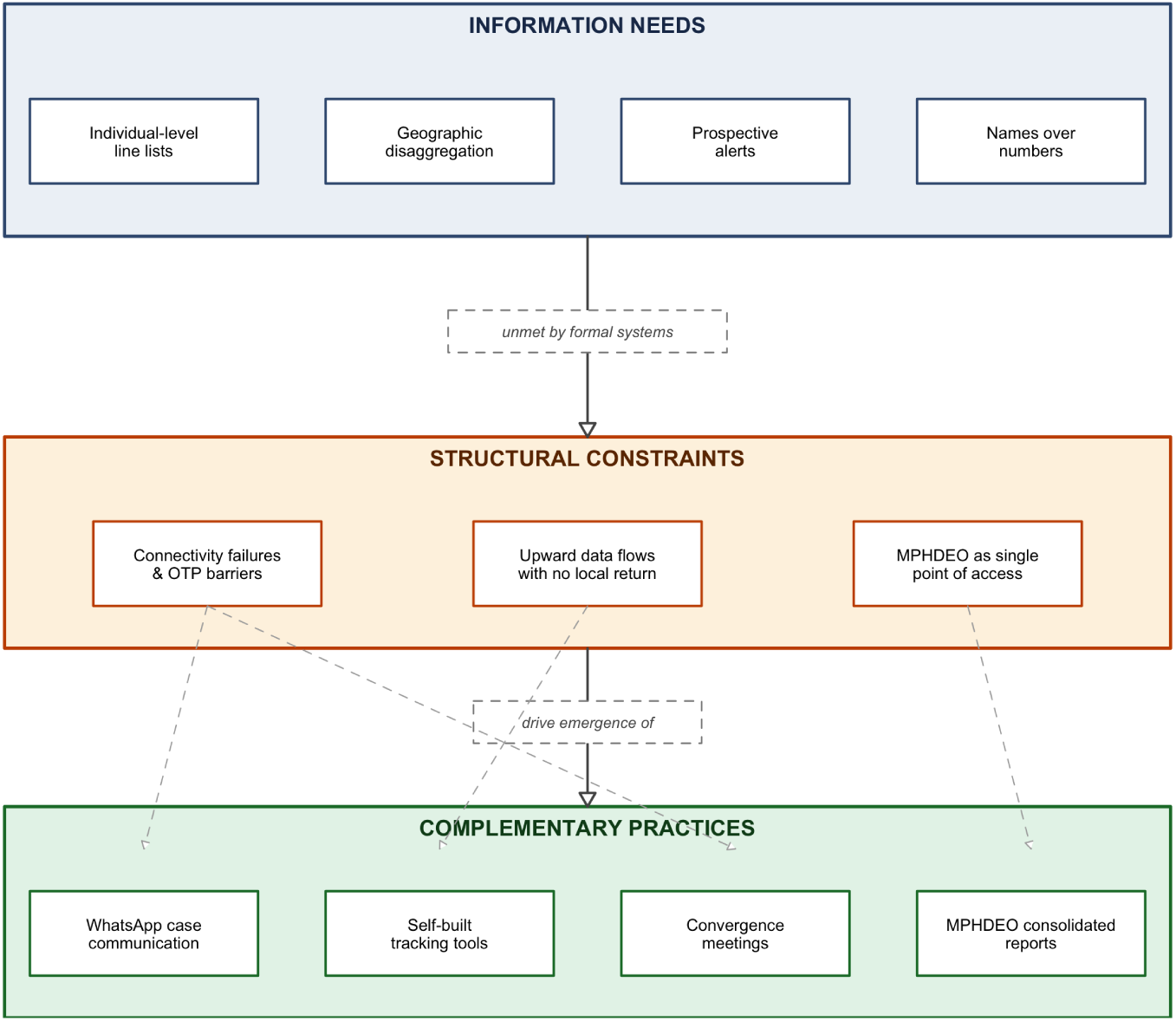
Concept map of findings: information needs that formal systems cannot deliver are explained by three structural constraints (connectivity failures, upward data flows, MPHDEO as single point of access), which in turn drive the emergence of four complementary information practices.

## 4 Discussion

### 4.1 Upward accountability and the data-decision gap in MCH systems

The finding that formal systems serve upward accountability rather than local decision support is consistent with the broader RHIS literature (1,2,16). What distinguishes the tribal India context is the layering of governance structures (PHC, ITDA, district, state) combined with infrastructure conditions (connectivity, terrain, seasonal access) that amplify the consequences of systems designed for connected, well-resourced settings. The OTP authentication requirement, a security measure appropriate for urban settings with reliable mobile coverage, functioned in this context as an access barrier that further widened the gap between data and decisions (17).

The medical officers in this study did not lack motivation to use data. Their expressed information needs were specific, clinically grounded, and operationally actionable. The constraint was structural: systems designed to satisfy supervisory data requirements at higher administrative levels did not return information to the level where clinical and programmatic decisions were made. This is consistent with what Braa and colleagues described as the “information paradox” in developing country health systems, where data collection increases without corresponding improvements in local data use (16).

### 4.2 The MPHDEO as gatekeeper: a governance problem

The concentration of portal access credentials in a single individual’s mobile phone represents a governance vulnerability that has not, to our knowledge, been previously described in the literature. It arises from the intersection of two policy decisions: (1) the shift to OTP-based authentication for the RCH Portal, and (2) the registration of institutional credentials to individual mobile numbers rather than institutional devices or multi-user authentication systems.

The MPHDEO occupies an ambiguous position. He is simultaneously a system vulnerability (single point of failure for data access) and a system adaptation (his consolidated reports are the most valued information product in the local MCH ecosystem). This dual role mirrors what the Open Data Institute describes as emergent data stewardship: individuals who, through institutional circumstance rather than formal mandate, become custodians of data access for their communities (10).

The policy implication is clear. Credential registration and authentication design for health information portals must be context-sensitive, not state-uniform. In settings where mobile network coverage is unreliable and institutional devices are unavailable, centralised OTP-based authentication creates access barriers that undermine the systems it is meant to secure.

### 4.3 Complementary information practices as local MCH data infrastructure

The four complementary practices identified in this study are not failures of the formal system. They are adaptive responses that reveal both the limitations of formal infrastructure and the informational priorities of local decision-makers. Framing them through the Open Data Institute’s concept of soft value exchanges highlights their function: they constitute flows of insights, knowledge, and feedback that operate outside formal data pipelines but serve essential MCH decision-making purposes (10).

This framing has parallels in Braa and colleagues’ concept of “networks of action” in sub-Saharan African health information systems, where local actors construct informal networks to compensate for formal system weaknesses (16). The practices identified here extend this concept by demonstrating a spectrum from complementary (convergence meetings operating alongside formal reporting) to substitutive (Google Sheets replacing portal functionality entirely). The position on this spectrum matters for intervention design: complementary practices can be strengthened without disrupting formal systems, while substitutive practices signal fundamental failures that require formal system reform.

### 4.4 The “names over numbers” imperative

The consistent demand for individual-level, geographically disaggregated information across all three PHCs reflects a fundamental mismatch between what health information systems report and what local MCH decision-making requires. Aggregate coverage statistics cannot drive individual case management for antenatal care, immunisation follow-up, or postnatal monitoring. This finding has direct design implications: any MCH tool intended for PHC-level use must provide drill-down from aggregate indicators to named beneficiary lists, organised by geographic unit, from the landing view (18,19).

This imperative is not unique to tribal settings, but the tribal context amplifies its urgency. Seasonal migration takes families away from registered villages, complicating follow-up. Naming conventions differ from portal assumptions, creating matching challenges. Small population denominators at the village level mean that a single missed case represents a substantial proportion of the eligible population. In this context, the distance between “5 pending” and “these 5 women in these villages” is the distance between information and action.

### 4.5 Implications for MCH digital health tools in tribal settings

Five implications emerge from these findings. First, new MCH digital health tools should be preceded by participatory diagnostics of the existing local information ecosystem, including complementary practices, to understand what the ecosystem already does before attempting to change it. Second, authentication and data access governance at the sub-district level requires reform. The MPHDEO access model is a systemic risk that demands policy attention at ITDA and state levels. Third, convergence meetings represent a model for cross-sectoral MCH data triangulation that could be formalised and extended. Fourth, the WhatsApp channel that medical officers already use and value should be integrated into, rather than replaced by, new tools. Fifth, offline functionality is not a feature but a prerequisite for any digital tool deployed in this context.

### 4.6 Limitations

This study has several limitations specific to its design and setting. Three PHCs in one ITDA jurisdiction provide depth but not breadth; findings may not transfer to better-resourced or non-tribal PHC contexts. Participant observation periods were limited to scheduled visits rather than continuous immersion. Self-reported accounts of data practices may reflect socially desirable responses, although the rapport established through the researcher’s community membership and the specificity of examples provided suggest candid engagement. The insider researcher effects were managed through independent coding by a second analyst (GJ), supervisor debriefs, and systematic attention to disconfirming evidence, but cannot be fully eliminated. The study captures diagnostic-phase findings before any intervention; post-intervention changes in practices and attitudes are reported elsewhere.

## 5 Conclusion

Local MCH decision-makers in tribal primary health centres are active information producers and knowledge managers, not passive data collectors. The gap between MCH data and local decisions is structural, embedded in authentication design, upward reporting logic, and sub-district governance arrangements, rather than a reflection of individual capacity or motivation.

The MPHDEO as sole data custodian across all PHCs in the jurisdiction represents an addressable governance vulnerability. Complementary information practices, including WhatsApp networks, self-built tracking tools, convergence meetings, and intermediary-consolidated reports, constitute latent MCH information infrastructure that digital health reform should formalise and strengthen rather than replace.

Future MCH health information system interventions in tribal settings should begin with participatory diagnostic of local information ecosystems, including the soft value exchanges that sustain local decision-making, before deploying new tools.

## Data Availability

All data produced in the present study are available upon reasonable request to the authors

## Declarations

## Ethics approval and consent to participate

Approved by the Institutional Ethics Committee, Sree Chitra Tirunal Institute for Medical Sciences and Technology (Protocol No. SCT/IEC/2047/MAY/2023). The CONSIDER statement for research involving Indigenous peoples informed the ethical framework (11).

## Consent for publication

Not applicable.

## Availability of data and materials

Interview transcripts are available from the corresponding author on reasonable request, subject to participant consent and ethics committee approval. The coding scheme and audit trail are available from AM.

## Competing interests

The authors declare no competing interests.

## Funding

The study did not receive any funding. However, it was a part of the PhD work of the AM (first author). He gratefully acknowledges the financial subsistence provided by the Science for Equity, Empowerment and Development (SEED) Division, Department of Science and Technology, Govt. of India, for the PhD work.

## Authors’ contributions

AM conceptualised the study, conducted field data collection, led the analysis, and drafted the manuscript. AM and GJ independently coded transcripts; discrepancies were resolved through deliberation. GJ also contributed to field data collection. BO, SKM, RN, and SS, as PHC Medical Officers and action research co-participants, provided contextual insights throughout the research process and reviewed the draft. RG and BS supervised the study design, qualitative methodology, and critically revised the manuscript. TB, EN, and SK served on the doctoral advisory committee and contributed to conceptualisation, review of findings, and structuring of the draft. AR contributed to field data collection. All authors reviewed and approved the final version.

## Acknowledgements

We thank the ANMs, ASHAs, and ITDA staff who participated in this study and shared their knowledge of local data systems.

## Notes

### Competing Interest Statement

The authors have declared no competing interest.

### Clinical Protocols

https://ctri.nic.in/Clinicaltrials/pmaindet2.php?EncHid=MTA0NDc4

### Author Declarations

The study received ethical approval from the Institutional Ethics Committee, Sree Chitra Tirunal Institute for Medical Sciences and Technology (Protocol No. SCT/IEC/2047/MAY/2023).

